# Algorithmic Ascertainment of Cause of Death from Longitudinal Real-World Medical Claims Data: Development and Validation

**DOI:** 10.64898/2026.08.18.26360606

**Authors:** Kyle W. McLean, Jason LaBonte, Kirsty Macaulay, Shahir Kassam-Adams

## Abstract

This study documents the derivation and validation of a deterministic algorithm for cause-of-death (COD) ascertainment from longitudinal real-world medical claims data, evaluated against an independent state-level death certificate file. Death certificates are the dominant reference standard in mortality research but carry well-documented limitations, including primary-cause error rates estimated at 20-40% across empirical studies. A matched analytic cohort of 216,382 individuals (Connecticut death records, 2017–2025, age 25 and above) was constructed after exclusion of mechanism-of-injury cases and removal of ill-defined symptom-code entries from both sources. Concordance between algorithmic and certificate-based COD was assessed through three complementary frameworks: age-stratified positive predictive value (PPV) at the ICD-10-CM chapter level under a full-set concordance scenario; mean absolute rank difference (MARD) for chapters identified by both sources; and analyses of breadth, depth, and code-level specificity of COD reporting. Chapter-level PPV was strongest for individuals aged 55 and above, with all estimates representing conservative lower bounds given the known error rate of the certificate reference standard. The algorithm consistently reported broader and more granular contributing cause profiles than the death certificate, with discordances directionally consistent with the well-documented tendency of certificates to under-report contributing conditions. These findings support the conclusion that algorithmic COD ascertainment from longitudinal claims data is a feasible and scalable alternative to certificate-based attribution and, at population scale, a principled methodology for characterising death certificate error rates beyond what small-sample chart review studies can achieve.

## Introduction

Cause of death (COD) is among the most consequential variables in health analytics. Understanding why patients die, not merely that they died, underpins a wide range of scientific and commercial endeavours: survival analysis and mortality endpoint adjudication in clinical trials and real-world evidence studies,^1^ risk stratification and care management in health care operations,^2^ life expectancy modelling in actuarial and financial services,^3,4^ pharmacovigilance signal detection,^5,6^ and population health surveillance.^7,8^ Without reliable COD attribution, researchers and analysts are left to work backward from incomplete signals, introducing systematic bias into downstream inference. As value-based care models increasingly tie reimbursement to outcomes, and as mortality indexes become embedded in risk adjustment methodologies, the demand for accurate, scalable COD data has grown substantially. Yet the sources from which COD is typically drawn each carry meaningful limitations that constrain their utility in large-scale data applications.

There are three principal sources of COD information in the United States: death certificates, self-reported records in obituaries or memorial notices, and retrospective clinical chart reviews of a deceased patient’s medical history.

- **Death certificates** are widely regarded as the gold standard for COD ascertainment. They provide universal coverage, with a COD recorded for virtually every registered death in the United States, and are collected through state vital statistics offices and, at the national level, through the Centers for Disease Control and Prevention’s National Death Index (NDI).^9^ Despite this, death certificates carry significant practical and accuracy limitations. Access is tightly restricted: only family members and parties with a documented legal interest (such as life insurers) may obtain individual certificates, and access to the NDI requires a formal application process that is both slow and expensive and restricted to non-commercial medical research and public health use cases. More critically, the accuracy of death certificates has been called into question by empirical evidence. A 2021 systematic review and meta-analysis of the global literature reported primary-cause error rates in the range of 20-40% ^10^; some institutional studies, in which death certificates were independently reviewed against clinical records, have found rates of incorrect underlying-cause designation as high as 60%.^11^ Because the certifying party may be a funeral director, medical examiner, or sheriff rather than an attending physician, and because even clinicians completing certificates frequently default to the most proximate diagnosis in a patient’s chart, systematic inaccuracies are common. ^12–14^ The importance of formal physician training in accurate certificate completion has been emphasised repeatedly in the literature, yet errors persist across institutional settings.^13,15^ This evidence substantially undermines the reliability of death certificate-based mortality data as an unqualified reference standard.
- **Self-reported COD information**, most commonly found in obituaries, offers a complementary signal. Obituary-sourced COD is publicly accessible, carries no use restrictions, and because it is not classified as medical data, requires no de-identification. Obituaries are also typically available more rapidly after death than any other structured source. However, a COD is reported in a substantial minority of obituaries, and the information provided is subject to social desirability bias: stigmatised causes such as suicide, drug overdose, and certain communicable diseases are generally omitted or obscured. When COD is referenced at all, it is often inferential rather than explicit - for example, a solicitation to donate to the Alzheimer’s Association in the decedent’s memory.^16,17^ Comorbid conditions that contributed to death are almost never reported.
- **Chart review** of the deceased’s medical history involves a reviewer working backward from the date of death through the patient’s medical record, physician notes, lab results, and hospital summaries, to identify the acute event that likely triggered death while also tracing chronic conditions that may have contributed. The strength of this approach is its accuracy and nuance: a trained reviewer can catch discrepancies, contextualise ambiguous documentation, and account for complex or multi-factorial cases that automated methods might miss. The major drawback is that it is resource-intensive, requiring significant time, clinical expertise, and cost per case, which makes it difficult to scale across large populations or datasets. It is also subject to reviewer variability and depends heavily on how complete and legible the underlying records are.

Taken together, these three sources exhibit meaningful shortfalls that limit the ability of researchers to curate accurate and comprehensive COD data, thus undermining outcomes derived from the resultant data. Since none of these sources is adequate at scale, studies require an alternative approach to COD database curation.

This convergence of limitations creates a clear methodological opportunity: a systematic, algorithm-driven approach to deriving COD from large-scale real-world clinical data (RWD) that codifies the logic of physician chart review into a reproducible, auditable pipeline capable of operating across populations of millions. A longitudinal record of diagnoses, procedures, medications, hospitalisations, and encounters reported in electronic health records (EHR) and insurance claims can be traced up to the point of death, mirroring the process a trained clinician would undertake during a formal chart review to assign COD. Analysing this data enables identification not only of a primary cause, but also of multiple comorbidities that meaningfully contributed to death - a richer characterisation than either death certificates or obituaries provide. Further, the use of a deterministic model allows complete traceability of any derived causes of death, which allows validation of such outputs for use as real-world evidence and in regulatory submissions. The remainder of this paper describes the development and validation of a deterministic algorithm designed for this purpose, its performance characteristics relative to death certificate attribution, and its implications for mortality research at scale.

## 1 Methods

### 1.1 Overview

The Veritas Cause of Death (COD) algorithm is a multi-stage computational pipeline that infers a primary and up to four secondary causes of death for individuals using longitudinal real-world medical claims data. The algorithm integrates confirmed death records with each patient’s antecedent clinical history, normalizes all clinical codes to a standardised cause-of-death taxonomy, computes a composite acuity-weighted score for each candidate diagnosis, and ranks those candidates to assign primary and secondary designations. A schematic overview is presented in Figure 1, with a breakdown of each stage provided in Table 1.

**Figure 1.**
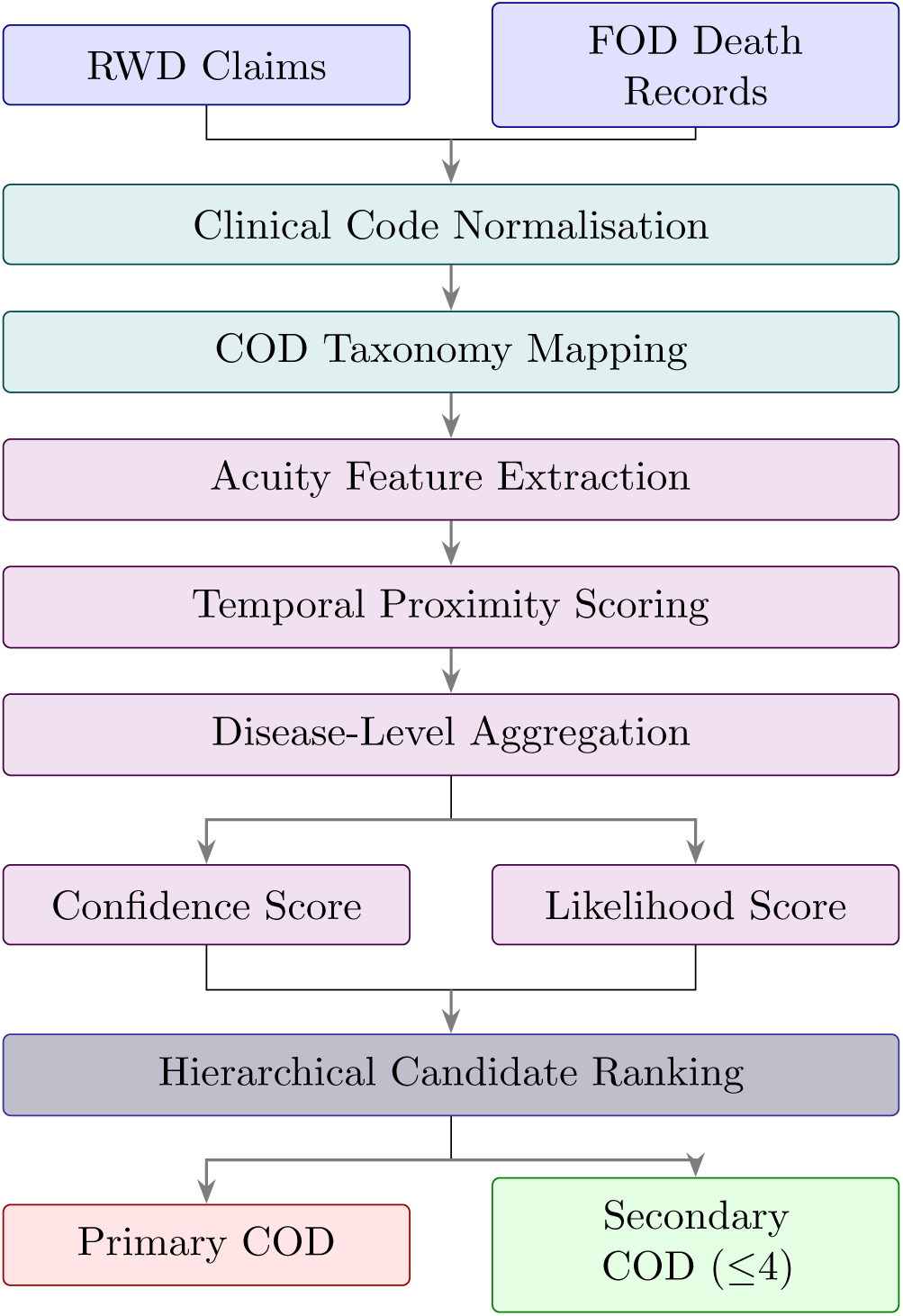
End-to-end pipeline for algorithmic cause-of-death ascertainment. RWD claims are linked to confirmed FOD death records, normalised through the UMLS Metathesaurus, and scored to yield one primary and up to four secondary causes of death.

**Table 1.** Stage-level breakdown of the cause-of-death algorithm.

| Stage | Details |
| --- | --- |
| RWD Medical Claims | ICD-9/10-CM · CPT · HCPCS · POS · Specialty · Modifiers |
| FOD Death Records | Confirmed date of death · Pseudonymous traceability ID |
| Clinical Code Normalisation | UMLS Metathesaurus (NLM) · CUI unification across all vocabularies |
| COD Taxonomy Mapping | CDC NCHS-113 hierarchy · Exact / Range / Prefix matching |
| Acuity Feature Extraction | Provider specialty · POS · Modifiers · Coefficients 1-5 · 90th-pct z-score |
| Temporal Proximity Scoring | Recency z-score (366-day linear decay) · Within-patient decile rank |
| Disease-Level Aggregation | 8 time windows: 7, 30, 60, 90, 180 d; 1, 2, 3 yr · Peak acuity · Admission count |
| Confidence Score | Frequency + Recency + 3-yr peak acuity |
| Likelihood Score | Recency + 180-d + 1-yr admission acuity |
| Hierarchical Candidate Ranking | Likelihood → Confidence → time-window tiebreakers · Both scores > 0.10 · Top 5 retained |

### 1.2 Study Population and Death Record Integration

Patient eligibility for COD analysis is determined by cross-referencing RWD medical claims against the Veritas Fact of Death (FOD) database. The FOD database is a proprietary, continuously updated repository of confirmed death records extracted, consolidated and deduplicated from over 45,000 sources and validated against United States vital records. Each death record is characterised by a confirmed date of death and associated demographic data, including date of birth, sex, geographic region (three-digit ZIP code), and state.

To protect patient privacy, all records are de-identified prior to analytical processing using an established Privacy Preserving token creation process.

Where multiple administrative sources provide conflicting demographic information for the same individual, a deterministic source-priority hierarchy is applied, taking into account source quality and diversity.

#### 1.2.1 Validation Cohort Definition

The validation cohort is derived from the intersection of Connecticut death certificate records and Veritas COD assignments, with four eligibility criteria applied uniformly to all individuals (Table 2 provides a reference summary of all ICD-10-CM chapters; chapters affected by criteria 2 and 4 are indicated therein):

1. **Death year restriction (2017-2025).** Only records with a confirmed death year between 2017 and 2025 inclusive are retained, ensuring that Veritas COD assignments are derived from years with complete and consistent claims coverage and that no partial calendar years are included at either endpoint of the observation window.
2. **Removal of individuals with V00-Y89 at any COD rank.** ICD-10-CM chapters V00-V99 (Transport accidents) and Y00-Y89 (Other external causes of morbidity and mortality) are excluded entirely from both sources for any individual carrying such a code at any rank position. The mechanism-of-injury codes in these chapters are structurally absent from longitudinal medical claims records, because claims data pertaining to external cause classification are redacted from the standard claims feeds ingested by the Veritas pipeline. Including these individuals would introduce a systematic and irreducible source of false negatives on the Veritas side, making the comparison unfair as an evaluation of the algorithm’s diagnostic inference capability.
3. **Minimum age at death (**≥ 25 **years).** Individuals aged below 25 at the time of death are excluded. The availability and completeness of longitudinal medical claims data for younger individuals is substantially lower than for older age groups: individuals under 25 typically have shorter insurance coverage histories and thinner claims records, providing an insufficient longitudinal signal for reliable algorithmic COD inference. Inclusion of these individuals would depress performance estimates in a manner that reflects data sparsity rather than algorithm quality.
4. **Exclusion of R00-R99 chapter entries from the comparison (individuals retained).** ICD-10-CM Chapter R00-R99 (Symptoms, signs, and abnormal clinical and laboratory findings, not elsewhere classified) is excluded as a *category* from the concordance analysis for both sources. Codes in this chapter are considered “ill-defined” and should not represent the underlying cause of death.^18,19^ Their presence in death certificates typically reflects incomplete certification rather than a meaningful biological cause. R-coded entries are stripped from both the CT and Veritas cause sets prior to analysis. *Individuals* whose records contain R-coded entries alongside other valid non-R causes are **retained** in the cohort; only the R-coded entries themselves are removed from the comparison.

**Table 2.** ICD-10-CM chapter classification used throughout this study. Chapters subject to exclusion under the eligibility criteria above are indicated: *^†^*entries removed from the comparison set, with individuals retained; *^‡^*individuals carrying codes in this chapter excluded from the cohort entirely.

| Chapter | Code range | Description |
| --- | --- | --- |
| I | A00–B99 | Certain infectious and parasitic diseases |
| II | C00–D49 | Neoplasms |
| III | D50–D89 | Diseases of the blood and blood-forming organs and certain disorders involving the immune mechanism |
| IV | E00–E89 | Endocrine, nutritional and metabolic diseases |
| V | F01–F99 | Mental, behavioral and neurodevelopmental disorders |
| VI | G00–G99 | Diseases of the nervous system |
| VII | H00–H59 | Diseases of the eye and adnexa |
| VIII | H60–H95 | Diseases of the ear and mastoid process |
| IX | I00–I99 | Diseases of the circulatory system |
| X | J00–J99 | Diseases of the respiratory system |
| XI | K00–K95 | Diseases of the digestive system |
| XII | L00–L99 | Diseases of the skin and subcutaneous tissue |
| XIII | M00–M99 | Diseases of the musculoskeletal system and connective tissue |
| XIV | N00–N99 | Diseases of the genitourinary system |
| XV | O00–O9A | Pregnancy, childbirth and the puerperium |
| XVI | P00–P96 | Certain conditions originating in the perinatal period |
| XVII | Q00–Q99 | Congenital malformations, deformations and chromosomal abnormalities |
| XVIII<sup>†</sup> | R00–R99 | Symptoms, signs and abnormal clinical and laboratory findings, not elsewhere classified |
| XIX | S00–T88 | Injury, poisoning and certain other consequences of external causes |
| XX<sup>‡</sup> | V00–Y89 | External causes of morbidity |
| XXI | Z00–Z99 | Factors influencing health status and contact with health services |
| XXII | U00–U85 | Codes for special purposes |

### 1.3 Real-World Data Ingestion

For each individual confirmed deceased in the FOD database, longitudinal medical claims records are retrieved from RWD sources. A set of medical *events* are curated for each individual, where each event represents a discrete clinical encounter and contains the following structured fields:

Primary and secondary **diagnosis codes** (ICD-9-CM or ICD-10-CM). The coding system is assigned from a source qualifier field where available, or inferred from the date of service relative to the ICD-10-CM implementation date of 1 October 2015.

- **Procedure codes** (CPT, HCPCS, or ICD-10-PCS).
- **Date of service**, expressed as days elapsed relative to the confirmed date of death (service day; negative values denote pre-death encounters).
- **Place of service** code, indicating the clinical setting of the encounter (e.g., inpatient hospital, emergency department, outpatient office).
- **Provider specialty**, recorded separately for the rendering, billing, referring, and facility provider roles.
- Up to four **procedure modifier** codes per encounter.
- **Diagnosis priority**, designating whether each code was recorded as the primary or a secondary diagnosis for that encounter.

The primary scoring model uses encounters occurring within 30 days prior to death. Encounters extending up to three years (1,095 days) before death are retained for supplementary recency and temporal context scoring.

### 1.4 Clinical Code Normalisation and COD Taxonomy Mapping

All clinical codes - regardless of the originating coding system - are unified through the Unified Medical Language System (UMLS) Metathesaurus (National Library of Medicine, annual release).^20^ The UMLS provides concept-level mappings across heterogeneous medical coding vocabularies, including ICD-9-CM, ICD-10-CM, ICD-10-PCS, CPT, HCPCS, SNOMED CT, and LOINC. Each clinical code is mapped to a UMLS Concept Unique Identifier (CUI), providing a coding-system-agnostic representation of the underlying clinical concept. Codes that resolve to more than one CUI are excluded to ensure unambiguous code-to-concept assignment.

CUIs are subsequently mapped to a structured cause-of-death taxonomy. This taxonomy is a curated extension of the Centers for Disease Control and Prevention (CDC) NCHS-113 cause-of-death classification. Each COD entry is organised within a four-level hierarchy: COD chapter, COD subchapter, NCHS-113 group, and ICD-10-CM code. Mapping is performed using a curated rule table that applies three matching strategies in descending order of specificity: (1) exact code match, (2) alphanumeric range match, and (3) string-prefix pattern match. Where multiple rules apply to the same code, the most specific rule takes precedence. Supplementary manual mappings are applied for clinical entities not yet represented in standard UMLS releases-for example, COVID-19 ICD-10-CM codes U07.0 and U07.1.

### 1.5 Acuity Score Construction

For each clinical encounter, a composite acuity score quantifies the severity and clinical intensity of that healthcare interaction. The score is derived from three encounter attributes, each mapped to an ordinal coefficient on a scale of 1 (very low acuity) to 5 (severe acuity): provider specialty (evaluated separately for rendering, billing, referring, and facility roles), place of service, and procedure modifier codes. The episode acuity score is the sum of all applicable coefficients:

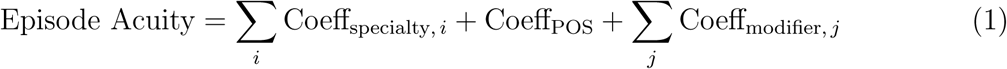

To normalise this score across the patient population and limit sensitivity to extreme outliers, a population-level reference value is established as the 90th percentile of episode acuity (*P*_90_) across all patients and encounters. The acuity z-score for each episode is then:

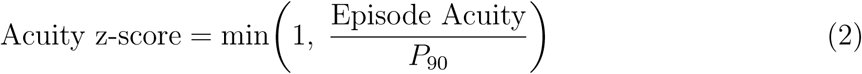

Scores exceeding the 90th percentile are capped at 1.0, yielding a bounded score on [0, 1].

### 1.6 Temporal Proximity Scoring

Two complementary metrics capture the temporal relationship between each clinical encounter and the date of death.

The **recency z-score** is a linear decay function that assigns maximum weight (1.0) to encounters on or immediately proximate to the date of death, declining to zero at 366 days before death:

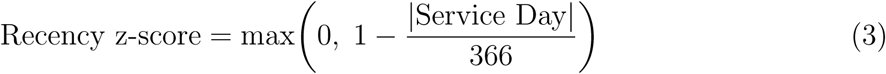

The **recency index** is a within-patient decile rank (1-10) assigned to each encounter based on its chronological position in that patient’s claims history, where decile 10 corresponds to the most recent encounters.

### 1.7 Disease-Level Aggregation

All encounters mapping to the same COD taxonomy code within a single patient are aggregated to produce a disease-level summary. Aggregation is performed across eight pre-specified time windows prior to death: 7, 30, 60, 90, and 180 days, and 1, 2, and 3 years. For each patient-disease pair, the following statistics are computed within each window:

- **Total events** - the number of distinct encounter dates associated with that disease.
- **Admissions count** - the number of distinct encounter dates on which the disease was recorded as the primary diagnosis.
- **Peak acuity z-score** - the maximum acuity z-score across all encounters for that disease in the window.
- **Peak admission acuity z-score** - the maximum acuity z-score restricted to encounters where the disease was the primary diagnosis.

### 1.8 Cause-of-Death Scoring

Two composite scores are computed per patient-disease pair to quantify the strength of evidence for each candidate cause of death.

#### 1.8.1 Confidence Score

The confidence score reflects the overall reliability of the evidence supporting a given cause of death, incorporating signal frequency, temporal recency, and peak clinical severity over the full three-year observation window:

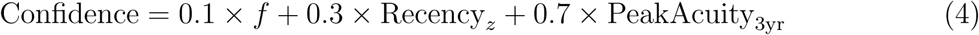

where *f* = 1.0 if the disease appears in more than two distinct encounters, and *f* = 0.7 otherwise. Confidence levels are assigned according to the thresholds in Table 3.

**Table 3.** Confidence level thresholds.

| Confidence Score | Assigned Level |
| --- | --- |
| $< 0.40$ | Low |
| $0.40\text{-}0.70$ | Medium |
| $> 0.70$ | High |

#### 1.8.2 Likelihood Score

The likelihood score quantifies the probability that a given disease is the cause of death, placing greatest weight on near-term clinical intensity:

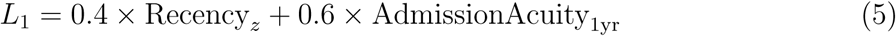

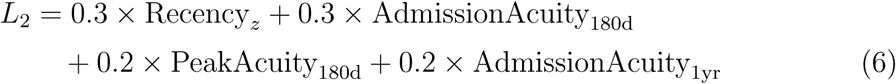

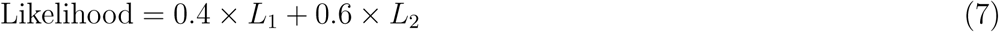

### 1.9 Primary and Secondary Cause of Death Designation

Candidate causes of death for each patient are ranked using a deterministic, hierarchical ordering scheme. The primary sort criterion is the likelihood score, with the confidence score as the first tiebreaker. Subsequent tiebreakers apply peak acuity z-scores, admissions counts, and total event counts sequentially across decreasing time windows from 7 days through to 3 years.

Up to five causes of death are retained per patient. The highest-ranked candidate (rank 1) is designated the **Primary Cause of Death**; ranks 2 through 5 are designated **Secondary Causes of Death**. Each retained cause is reported with its ICD-10-CM code, plain-language description, NCHS-113 group, subchapter and chapter classifications, likelihood and confidence scores, and ordinal rank designation.

## 2 Statistical Evaluation Framework

Agreement between Veritas algorithm output and Connecticut state death certificate records is assessed at the ICD-10-CM chapter level. This taxonomic resolution was selected to ensure that the comparison is clinically meaningful-chapters correspond to coherent disease domains whose epidemiological and clinical properties are well-characterised-while providing sufficient abstraction to allow reliable characterisation of PPV across subgroups. Finer resolutions (subchapter, NCHS-113 group, ICD-10-CM code) introduce increasing code-space cardinality and inter-rater variability that are unrelated to algorithm quality, and would produce underpowered per-cell estimates for most disease-age combinations in this cohort.

### 2.1 Full-Set Concordance

Concordance is evaluated under the full-set scenario. For each individual *i*, let V*_i_* denote the set of ICD-10-CM chapters present in the Veritas-assigned causes and R*_i_* the set of chapters present in the CT-recorded causes. To prevent reporting-breadth differences from inflating false-positive counts, the Veritas set is truncated to the same cardinality as the reference set prior to analysis: the top *k* Veritas-ranked causes are retained, where *k* = |R*_i_*|. Confusion matrix elements are accumulated over all chapter categories appearing in either set:

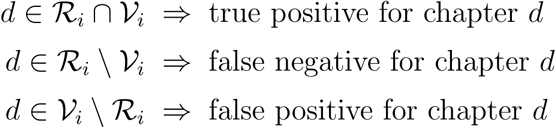

The full-set scenario is adopted as the primary evaluation framework because it appropriately accommodates the known imperfection of the death certificate as a reference standard. Given empirical evidence that 20-40% of death certificates carry an incorrect primary cause designation,^10,11^ penalising Veritas for cases where the correct chapter is present in the ranked list but not designated primary would conflate algorithm ranking behaviour with reference standard error. Evaluating whether the algorithm identifies the correct constellation of contributing chapters - irrespective of rank priority - is the operationally relevant question for the majority of downstream use cases.

### 2.2 Positive Predictive Value

For each chapter *d*, the four confusion matrix elements over the *N* individuals in the analytic cohort are:

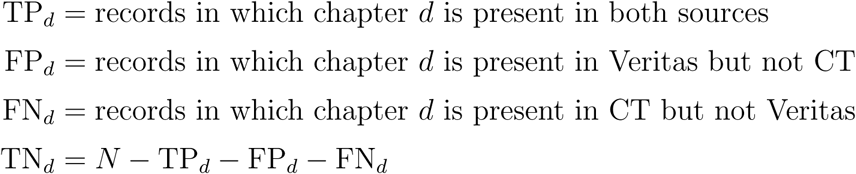

Because a single individual may contribute to the chapter-level counts of multiple chapters (multi-coded individuals contribute wherever a chapter is present in their Veritas set), the 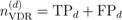 across all chapters will generally exceed *N* .

Positive Predictive Value is:

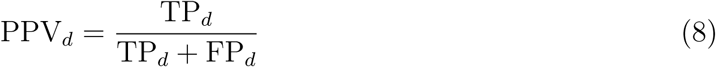

PPV quantifies the reliability of Veritas assignments for chapter *d*: the proportion of algorithm- positive records confirmed by the reference standard. Low PPV indicates systematic over-assignment, with direct implications for the inflation of disease-specific mortality burden in downstream analyses.

### 2.3 Statistical Power and Confidence Interval Framework

PPV estimates for individual chapter-by-age-band cells are accompanied by 95% Wilson score confidence intervals.^21^ The Wilson interval is preferred over the Wald interval because it maintains nominal coverage for small samples and extreme proportions, conditions that arise in chapter-level COD data where some strata contain few observations.

For a PPV estimate 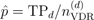 with 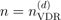, the Wilson 95% confidence interval is:

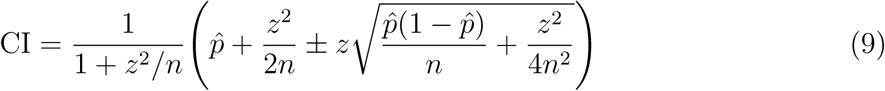

where *z* = 1.96 for a 95% interval. A minimum-power threshold is applied: the Wilson interval half-width is maximised at *p*^ = 0.5, giving an approximate half-width of z/(2√*n*). Setting a maximum acceptable half-width of 5 percentage points:

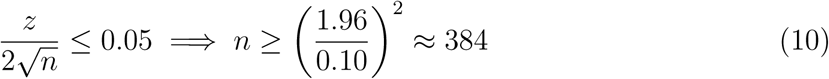

Cells with 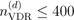 are excluded from reported PPV estimates and hatched in heatmap visualisations. All reported PPV values are therefore accompanied by a 95% CI with a half-width of at most 5 percentage points.

### 2.4 Macro and Micro Averaging

Per-chapter PPV values are aggregated across all chapters meeting the power threshold within a given age band using two approaches.

**Macro-average** assigns equal weight to each chapter and is restricted to chapters meeting the power threshold, denoted D*^∗^*. Excluding underpowered cells is necessary here because the unweighted mean treats each point estimate equally; a noisy PPV estimate from a cell with *n* = 10 would carry the same influence as one from a cell with *n* = 2,000:

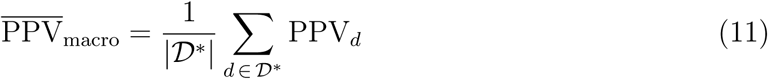

**Micro-average** pools raw TP and FP counts across *all* chapters before computing PPV, with no power-threshold restriction:

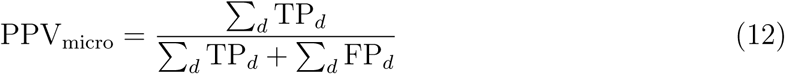

Because micro-averaging operates on pooled counts rather than point estimates, underpowered chapters contribute proportionally to their sample size and do not distort the aggregate. No threshold restriction is therefore needed or appropriate. Divergence between macro and micro estimates indicates that algorithm performance differs across high- and low-prevalence chapters. Both are reported as standard practice for evaluation on class-imbalanced problems.^22^

### 2.5 Order Analysis

The PPV analysis establishes whether the two sources identify the same ICD-10 chapters. The order analysis addresses the complementary question: for chapters that *are* identified by both sources, do the two sources assign consistent rank positions?

#### 2.5.1 Set Overlap (Jaccard)

Cause of death is not a single-valued data point: each decedent may have multiple contributing conditions recorded across both sources at varying rank positions. Treating COD as a set rather than a scalar therefore provides more clinical insight. Because death certificates are known to carry substantial inaccuracies in primary-cause designation,^10,11^ a metric that evaluates the full set of identified chapters - without privileging any single rank - provides a fairer axis of comparison between sources. Set overlap measured by the Jaccard coefficient focuses on what the two sources agree upon in terms of *which* disease domains are present, sidestepping the primary-versus-contributing distinction that is most susceptible to reference standard error. This simultaneously yields insight into the detection capability of the Veritas algorithm and surfaces patterns in death certificate reporting drawbacks documented in the literature: chapters where Jaccard is systematically low despite apparent clinical relevance are candidates for deeper examination of certificate under-reporting.

Per-record Jaccard similarity at the chapter level is computed across the full analytic cohort:

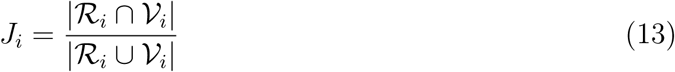

where V*_i_* is the Veritas set truncated to |R*_i_*| causes. The population mean *̄J* quantifies unconditional set overlap, defined by:

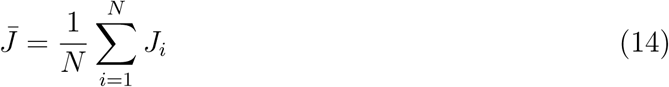

where N is the number of individuals in the cohort.

#### 2.5.2 Mean Absolute Rank Difference

Once set overlap is established, a further question arises: for the chapters that *are* identified by both sources, do the two sources assign them consistent rank positions? MARD addresses this conditional question. It is meaningful only after confirming non-trivial Jaccard similarity, because rank comparison is undefined when the chapter intersection is empty, and uninformative when overlap is negligible. Together, Jaccard and MARD decompose source agreement into two independent axes - *what* is detected and *how* it is prioritised - which supports a more precise diagnosis of where algorithm behaviour and reference standard limitations diverge.

For each individual *i*, let S*_i_*= R*_i_* ∩ V*_i_*be the intersection of agreed chapters. Analysis is restricted to individuals with |S*_i_*| ≥ 1. For each chapter *d* ∈ S*_i_*, let *r* (*d, i*) and *r* (*d, i*) denote the rank positions assigned by CT and Veritas respectively. The absolute rank difference is:

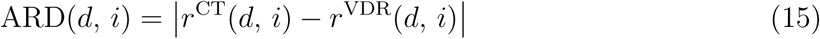

The **Mean Absolute Rank Difference** (MARD) is the mean of ARD over all chapter- individual pairs in the intersection:

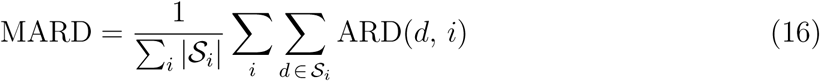

A MARD of 0 indicates perfect rank agreement for all shared chapters; a MARD of 1 indicates shared chapters are on average displaced by one rank position. Per-chapter MARD values identify which disease categories show the greatest rank instability between sources.

### 2.6 Completeness and Granularity of COD Reporting

Beyond concordance with the death certificate reference standard, three supplementary analyses characterise the structural properties of the two data sources as COD reporting instruments. These analyses do not evaluate algorithmic accuracy against a reference standard; rather, they quantify the degree to which a multi-year longitudinal claims record captures additional clinical signal relative to a point-in-time death certificate. Differences that systematically favour Veritas are expected *a priori* given the asymmetry in observational scope, and are interpreted as evidence of the richer characterisation afforded by longitudinal RWD.

#### 2.6.1 Breadth of Reporting

Breadth is defined as the number of distinct cause-of-death categories reported per individual. For each individual in the analytic cohort, the number of distinct causes of death present in the Veritas output 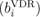 is compared with the number present in the CT death certificate record 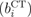. Veritas is considered to report equal or greater breadth when 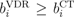. The comparison is additionally stratified across four taxonomic levels - raw ICD-10-CM code, subchapter, NCHS-113 group, and chapter - to assess whether breadth differences are consistent across classification granularities.

#### 2.6.2 Depth of Reporting

Depth quantifies the number of distinct ICD-10-CM codes reported within a matched cause-of-death category. The analysis is restricted to individuals for whom both sources report at least one cause in the same NCHS-113 group. Within each such matched group, the set of distinct ICD-10-CM codes contributed by each source is collected per individual. Parent codes that are strict prefixes of longer codes within the same set are removed prior to counting, ensuring codes are not double-counted via substring containment. The signed depth difference per individual is:

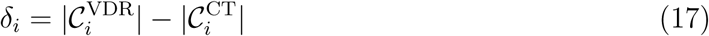

where 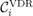 and 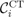 denote the deduplicated sets of ICD-10-CM codes within the matched NCHS-113 group for individual *i*. Statistical significance of the directional depth difference is assessed using a two-sided Wilcoxon signed-rank test applied to all *δ_i_* ≠ 0.

#### 2.6.3 Specificity of Reporting

Specificity is assessed at the ICD-10-CM code level. Among all code pairs where both sources report a code sharing the same three-character alphanumeric stem, the fourth character of each source’s code is examined. A fourth character of ‘9’ denotes an unspecified or not-otherwise-classified subcategory, while any other fourth character indicates a more specific classification. For each matched stem-pair, Veritas is classified as more specific when its fourth character is not ‘9’ and the CT fourth character is ‘9’, equal when both fall in the same category, and less specific in the converse case. Aggregate counts of these three outcomes are reported.

## 3 Validation Results

### 3.1 Study Population

Algorithm outputs were evaluated against cause-of-death records from the Connecticut state (CT) death certificate file, obtained from the Connecticut Department of Public Health. As discussed in Section 1, death certificate cause-of-death reporting has significant limitations; all concordance statistics reported here should be interpreted as lower bounds on true algorithm performance (see Section 4.5).

The analytic cohort was derived through deterministic record linkage and application of the four eligibility criteria described in Section 1.2.1. The derivation is summarised in Table 4.

**Table 4.** Analytic cohort derivation. Selection criteria applied: death years 2017-2025; no V00-Y89 chapter at any COD rank; age ≥ 25 at death; R00-R99 chapter entries excluded from comparison with individuals retained.

| Derivation Step | $N$ | % |
| --- | --- | --- |
| CT death certificate universe (distinct decedents with $\geq 1$ COD code) | 1,091,682 | — |
| Linked to Veritas RWD | 272,758 | 25.0% |
| Individuals with Veritas COD assignment | 247,904 | 22.7% |
| **After selection criteria applied (analytic cohort)** | **216,382** | **19.8%** |

The analytic cohort is stratified into six age bands at death: 25-34, 35-44, 45-54, 55-64, 65-74, and 75 and over. The distribution is markedly right-skewed, with the majority of individuals in the 65-74 and 75+ bands, reflecting the age distribution of mortality in the United States general population. This skew is not corrected by class reweighting or resampling, as the observed distribution represents the epidemiology of mortality in the matched population and the operational context in which the Veritas algorithm is deployed. The power threshold (Section 2.3) handles underpowered strata: cells that do not meet the *n*_VDR_ *>* 400 criterion are excluded from aggregated statistics and hatched in visualisations.

To verify that the cohort age-at-death distribution is representative of the national mortality pattern, the observed distribution was compared against CDC national mortality age distribution benchmarks for the corresponding years (2018-2024). The comparison is presented in Figure 2.

**Figure 2.**
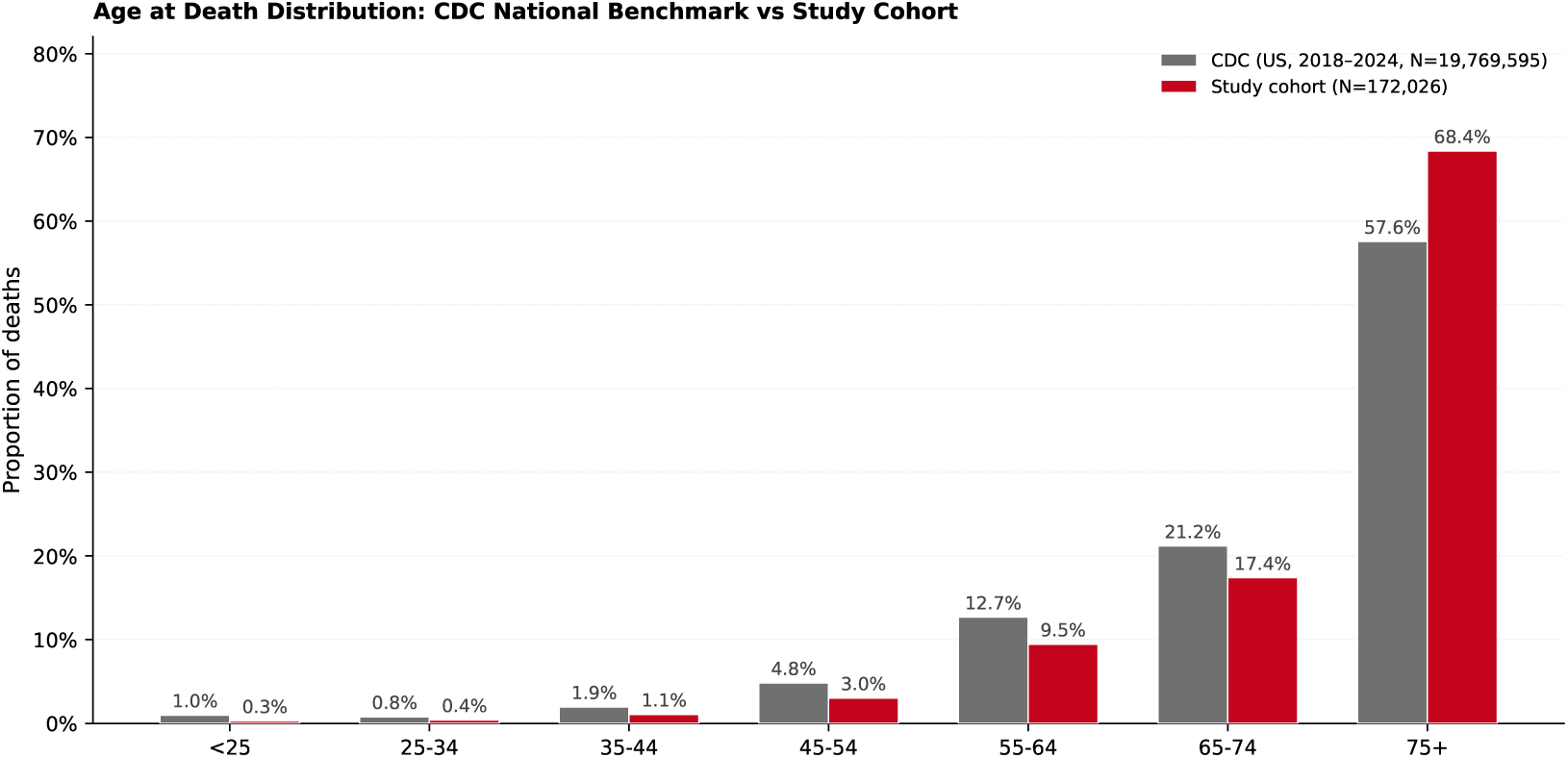
Age-at-death distribution for sample of the analytic cohort (*N_sample_* = 172,026) compared against CDC national mortality age distribution benchmarks for 2018-2024. PSI = 0.06, below the conventional threshold of 0.10 for negligible distributional shift. Cohen’s *w* = 0.229; interpretation is discussed in the text.

Two complementary measures of distributional stability were computed. The Population Stability Index (PSI) was 0.06, below the widely used threshold of 0.10 that demarcates negligible from moderate distributional shift. Cohen’s *w*, derived as 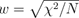. was 0.229, placing it in the small-to-medium range under Cohen’s conventional benchmarks (small *<* 0.10, medium *<* 0.30, large ≥ 0.50).

Cohen’s *w* has a structural property relevant to the interpretation of this value. Each age bin contributes a term 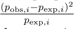 to the underlying chi-square statistic, meaning bins with small expected national proportions - the younger age strata, where few deaths occur at the population level - receive larger weight per unit of absolute deviation than the older age strata with large expected proportions. In this study the analytically and clinically important strata are the 55 and above age bands, which account for the majority of cohort individuals, the majority of powered chapter-by-age-band cells, and the most reliable PPV estimates. These correspond precisely to the high-frequency bins that are weighted least by Cohen’s *w*. The value of 0.229 therefore reflects disproportionate sensitivity to deviations in low-frequency young age strata - strata that are treated as exploratory in this study given their underpowered cell estimates and wider confidence intervals. The PSI of 0.06 provides a complementary summary that is not subject to this amplification and indicates that the cohort age distribution is consistent with the national mortality pattern for the purposes of this study.

### 3.2 Metric 1 - Age-Stratified Chapter-Level PPV

#### 3.2.1 Design

PPV was computed at the ICD-10-CM chapter level under the full-set concordance scenario (Section 2.1), stratified by six age bands at death. The ICD-10-CM chapter is the appropriate level of taxonomic resolution for this primary analysis: it provides clinically meaningful disease domain groupings - neoplasms, circulatory disease, respiratory disease, and so forth - that correspond to coherent mortality risk categories relevant to health analytics applications, while aggregating sufficiently to yield powered cell estimates across the age-band stratification. Evaluation at finer taxonomic levels (subchapter or individual code) would substantially increase the proportion of underpowered cells without providing proportional gains in interpretive value.

The Veritas cause set is truncated to the same cardinality as the CT cause set for each individual (top *k* causes, *k* = |R*_i_*|), ensuring that differences in reporting breadth do not inflate false-positive counts. R00-R99 entries are excluded from both sets prior to all computations as described in Section 1.2.1.

#### 3.2.2 Results

Chapter-level PPV by age band is presented in the heatmap in Figure 3. Macro- and micro-average PPV per age band are shown in Figure 4. Complete per-cell PPV values, Wilson 95% confidence intervals, and *n*_VDR_ counts are provided in the accompanying supplementary data.

**Figure 3.**
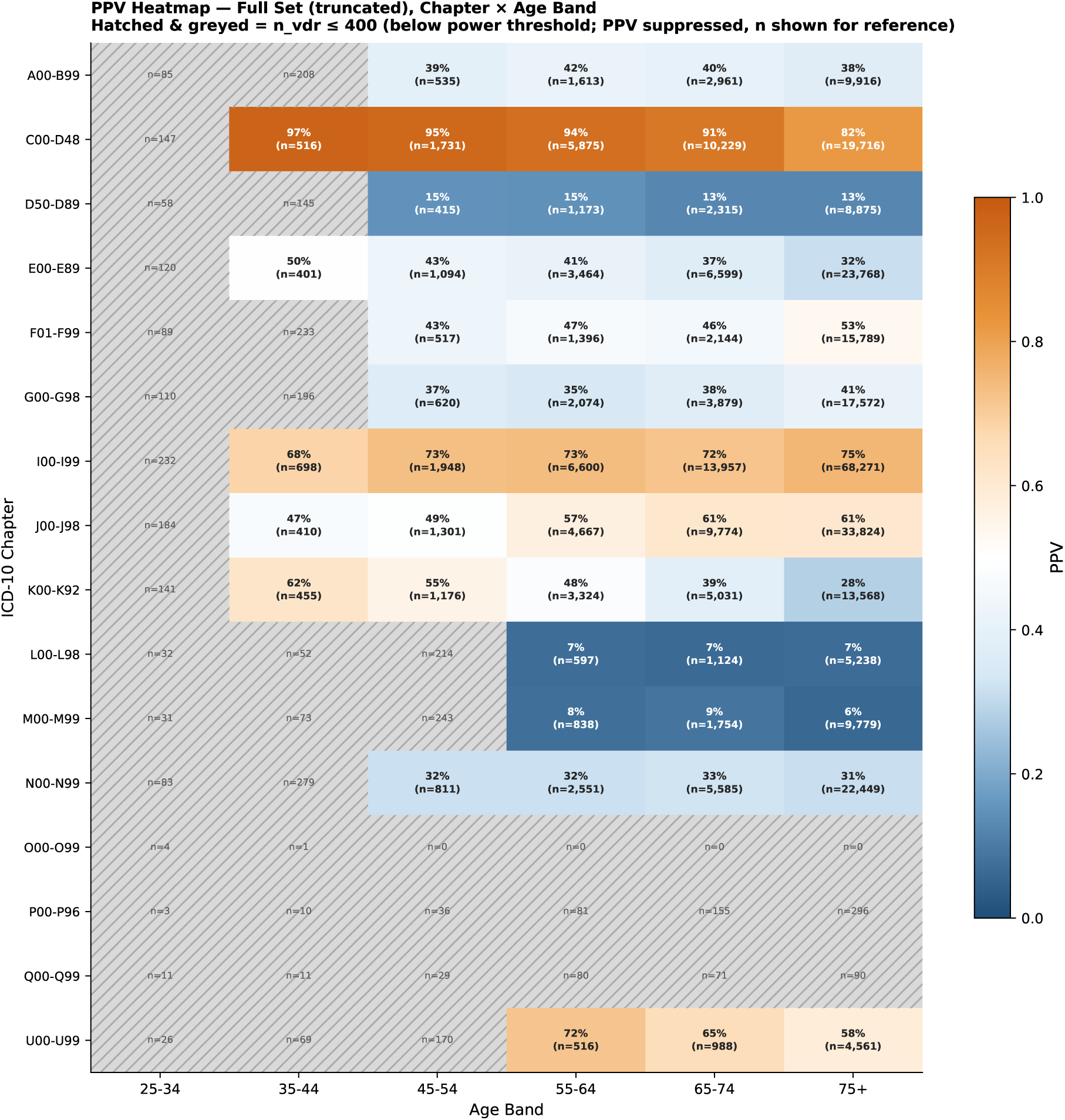
Chapter-level PPV heatmap under the full-set concordance scenario, stratified by age band at death (*N* = 216,382). Each cell reports PPV for a given ICD-10-CM chapter and age band. Hatched cells did not meet the minimum power threshold (*n*_VDR_ ≤ 400) and are excluded from macro-average calculations. The Veritas set is truncated to the CT set size per individual prior to analysis. R00-R99 and V00-Y89 chapter entries are excluded.

**Figure 4.**
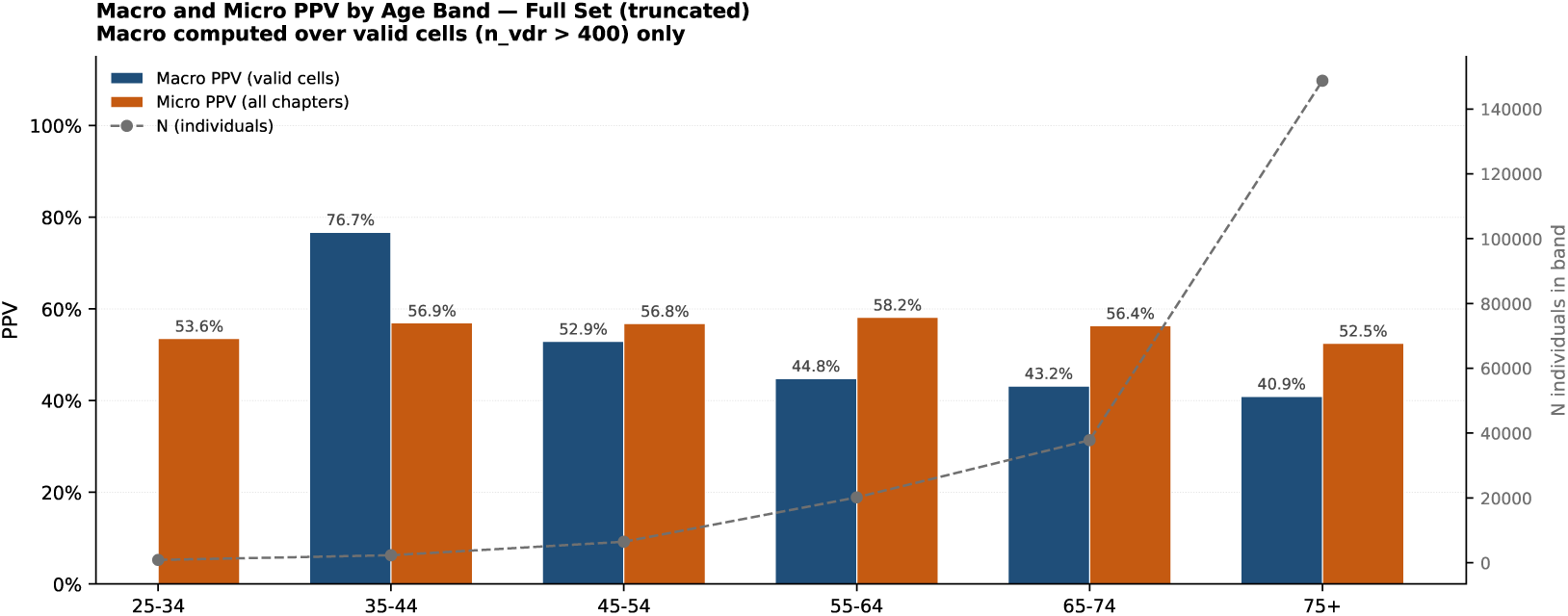
Macro-average and micro-average PPV by age band under the full-set concordance scenario. Macro-average is the unweighted mean of per-chapter PPV restricted to cells meeting the power threshold (*n*_VDR_ *>* 400). Micro-average is the pooled TP/(TP+FP) across all chapters unconditionally; no power-threshold restriction is applied.

The heatmap reveals age-dependent heterogeneity in concordance: performance is consistently strongest for individuals aged 55 and above, where both RWD claims density and per-cell sample sizes are greatest. PPV estimates for the 25-34 and 35-44 bands should be treated with greater caution, as fewer chapter-by-age-band cells meet the reporting threshold and confidence intervals are correspondingly wider.

All PPV estimates should be interpreted as lower bounds on true algorithm accuracy. Because the reference standard itself carries an estimated 20-40% primary-cause error rate,^10,11^ some proportion of the apparent false positives reflect correct Veritas assignments that were absent from or incorrect on the death certificate, rather than algorithm errors (see Section 4.5 for extended discussion).

### 3.3 Metric 2 - Order Analysis

The order analysis was conducted on the full analytic cohort (*N* = 216,382). MARD was computed conditional on individuals with at least one shared chapter between sources (|S*_i_*| ≥ 1). Results are presented in Figures 5-7.

**Figure 5.**
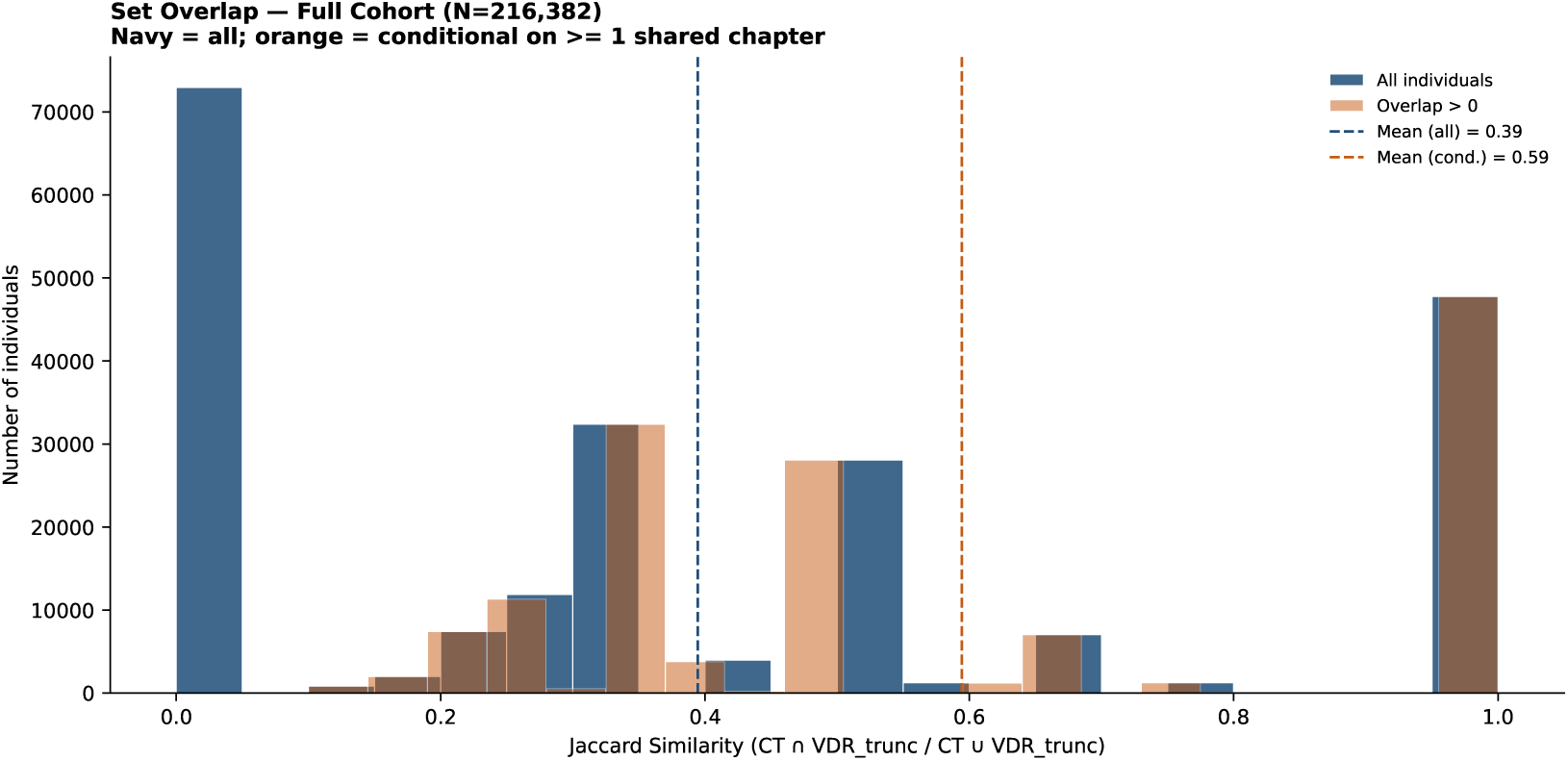
Distribution of per-record Jaccard similarity at the ICD-10-CM chapter level across the full analytic cohort (*N* = 216,382), quantifying unconditional set overlap between sources. Jaccard = 0 indicates no shared chapters between sources for that individual; Jaccard = 1 indicates complete set agreement.

**Figure 6.**
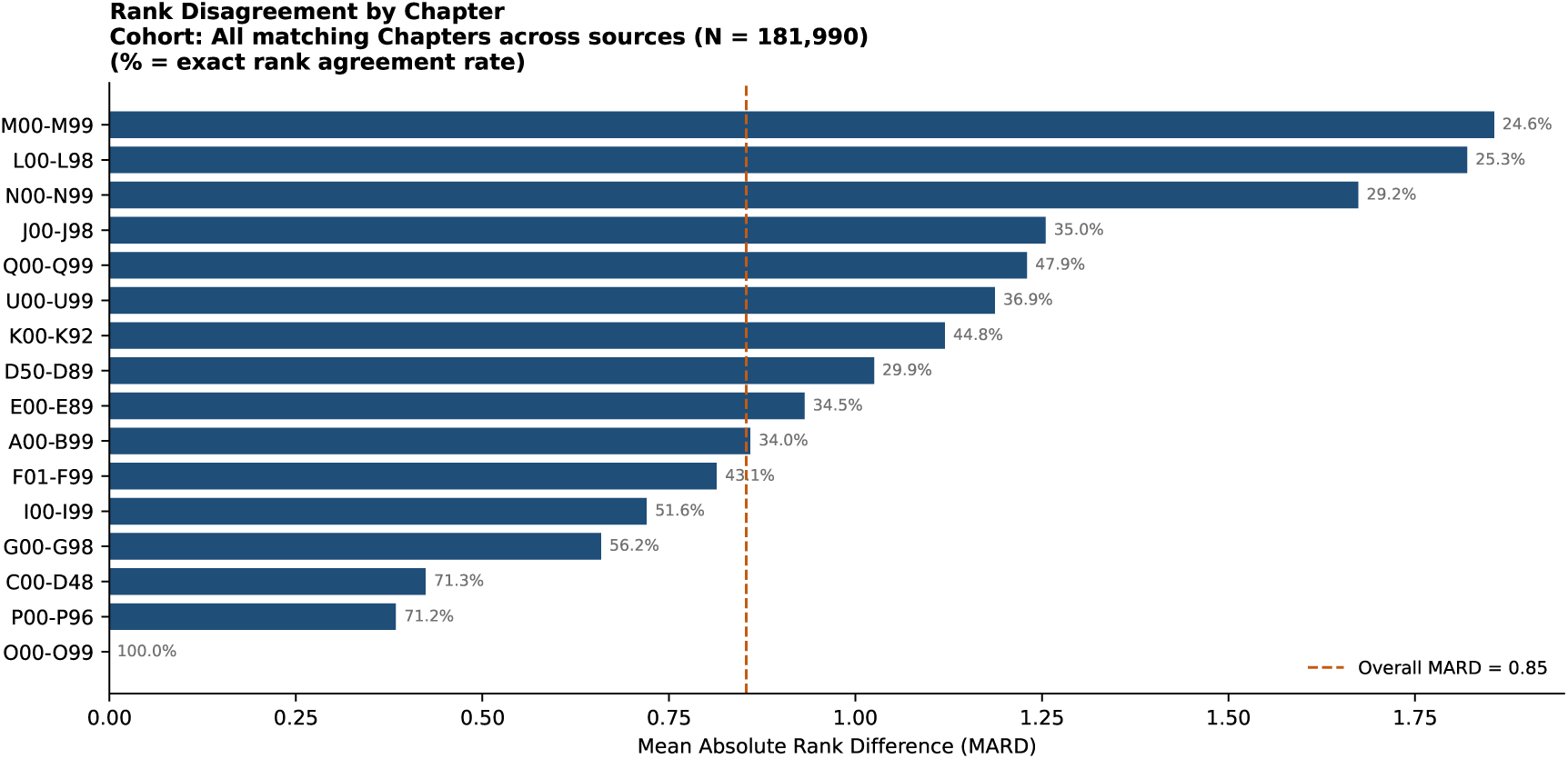
Per-chapter mean absolute rank difference (MARD), restricted to individuals with ≥ 1 shared chapter (|S*_i_*| ≥ 1). Higher MARD indicates greater rank instability for a chapter between the two sources. The dashed vertical line indicates the overall MARD across all shared chapter-individual pairs.

**Figure 7.**
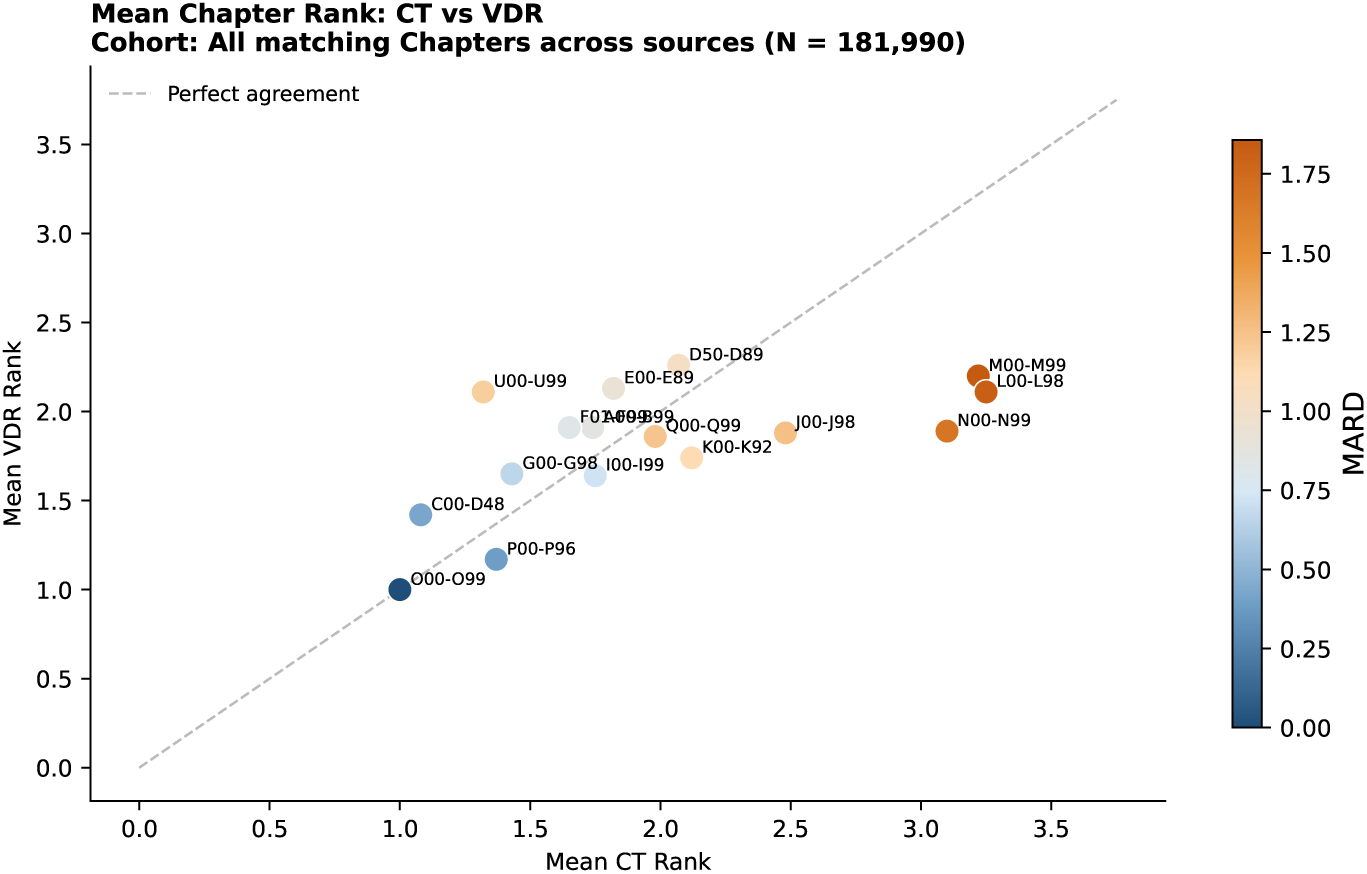
Mean CT rank versus mean Veritas rank for each ICD-10-CM chapter, among individuals with ≥ 1 shared chapter. Points on the diagonal indicate perfect average rank agreement between sources; colour encodes per-chapter MARD. Chapters above the diagonal are ranked higher on average by Veritas than by CT; chapters below are ranked higher by CT.

Figure 5 characterises the frequency with which the two sources identify the same chapter constellation; Figure 6 characterises, conditional on chapter agreement, whether the two sources assign the chapter a consistent rank position. These measures address complementary aspects of source concordance. High Jaccard with low MARD indicates that the sources agree both on which chapters are present and on their relative priority. High Jaccard with elevated MARD indicates that the sources detect the same disease domains but diverge in how they prioritise them - a pattern that reflects the well-documented difficulty in reliably designating an underlying versus contributing cause when multiple chronic conditions are present simultaneously.

Per-chapter MARD values (Figure 6) identify disease categories with the greatest rank instability between sources. Chapters with consistently high MARD represent cases where the Veritas algorithm and the certifying physician agree on the presence of a condition but assign it different priority levels. Interpreting elevated MARD requires acknowledging that rank disagreement may reflect reference standard error - the certifying physician’s primary-cause designation is known to be unreliable in a substantial fraction of cases^12,13^ - rather than solely an algorithm calibration issue. Figure 7 provides a complementary view of the same data, showing the relationship between mean CT and mean Veritas rank positions across chapters and identifying systematic directional biases.

### 3.4 Metric 3 - Completeness and Granularity of COD Reporting

#### 3.4.1 Breadth of Reporting

Across the analytic cohort, Veritas reported equal or more distinct NCHS-113 cause-of-death groups than the Connecticut death certificate in 166,137 of 216,382 individuals for whom both sources contributed at least one reportable category (76.8%); the CT death certificate reported more distinct groups in the remaining 50,245 individuals (23.2%). Figure 8 presents the aggregate breakdown. Figure 9 shows the frequency distribution of the number of COD categories reported per individual at four taxonomic levels (ICD-10-CM chapter, subchapter, NCHS-113 group, and raw ICD-10-CM code). Across all four levels, Veritas produces a broader distribution, with a higher proportion of individuals assigned multiple contributing causes.

**Figure 8.**
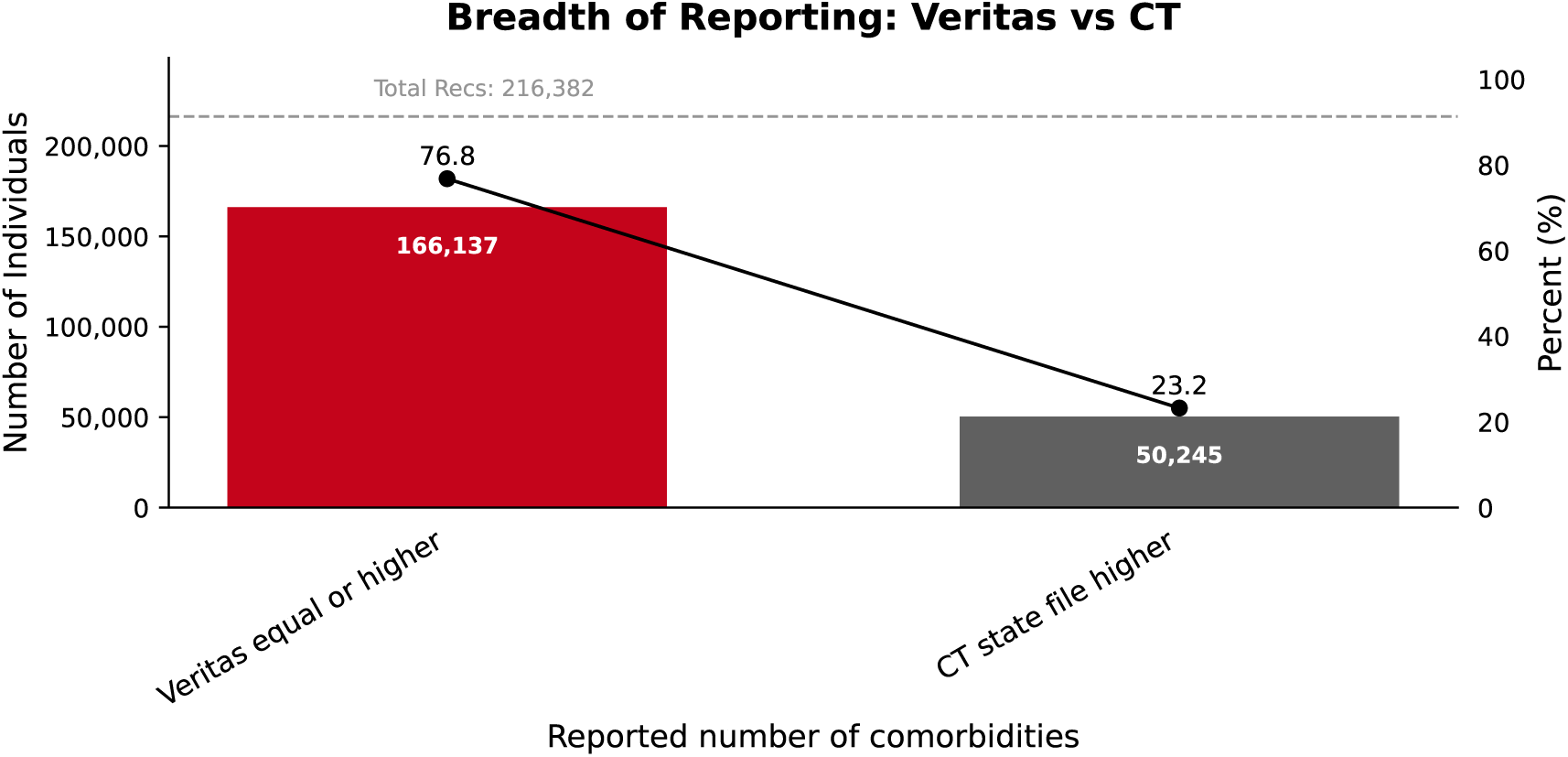
Breadth of cause-of-death reporting at the NCHS-113 group level (*n* = 216,382 individuals with at least one reportable category in both sources). Veritas reported equal or greater breadth than the CT death certificate in 76.8% of individuals.

**Figure 9.**
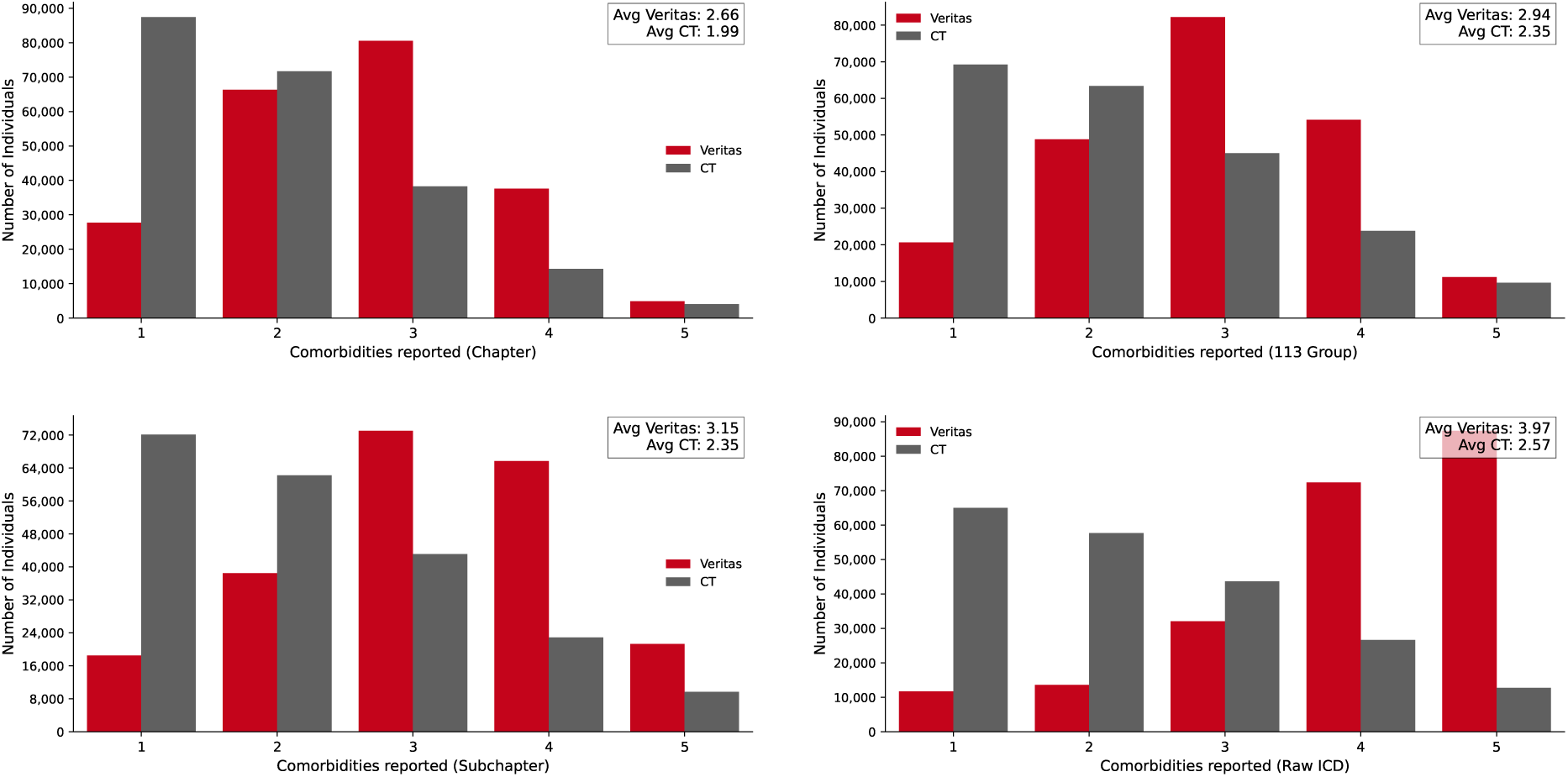
Frequency distribution of the number of distinct cause-of-death categories reported per individual, shown at four taxonomic levels: ICD-10-CM chapter (top left), NCHS-113 group (top right), subchapter (bottom left), and raw ICD-10-CM code (bottom right). Veritas distributions extend to higher counts at all levels, reflecting the broader clinical signal captured by multi-year longitudinal claims data relative to a single death certificate.

#### 3.4.2 Depth of Reporting

Among individuals with at least one matched NCHS-113 group between sources, 57,810 records contributed non-zero depth differences (*δ_i_* ≠ 0). Of these, 77.7% showed Veritas reporting a greater number of distinct ICD-10-CM codes within the matched category than the CT death certificate. The Wilcoxon signed-rank test confirmed the direction and magnitude of this difference as statistically significant (statistic = 1.32 × 10^9^; *p* ≈ 0). Results are presented in Figure 10.

**Figure 10.**
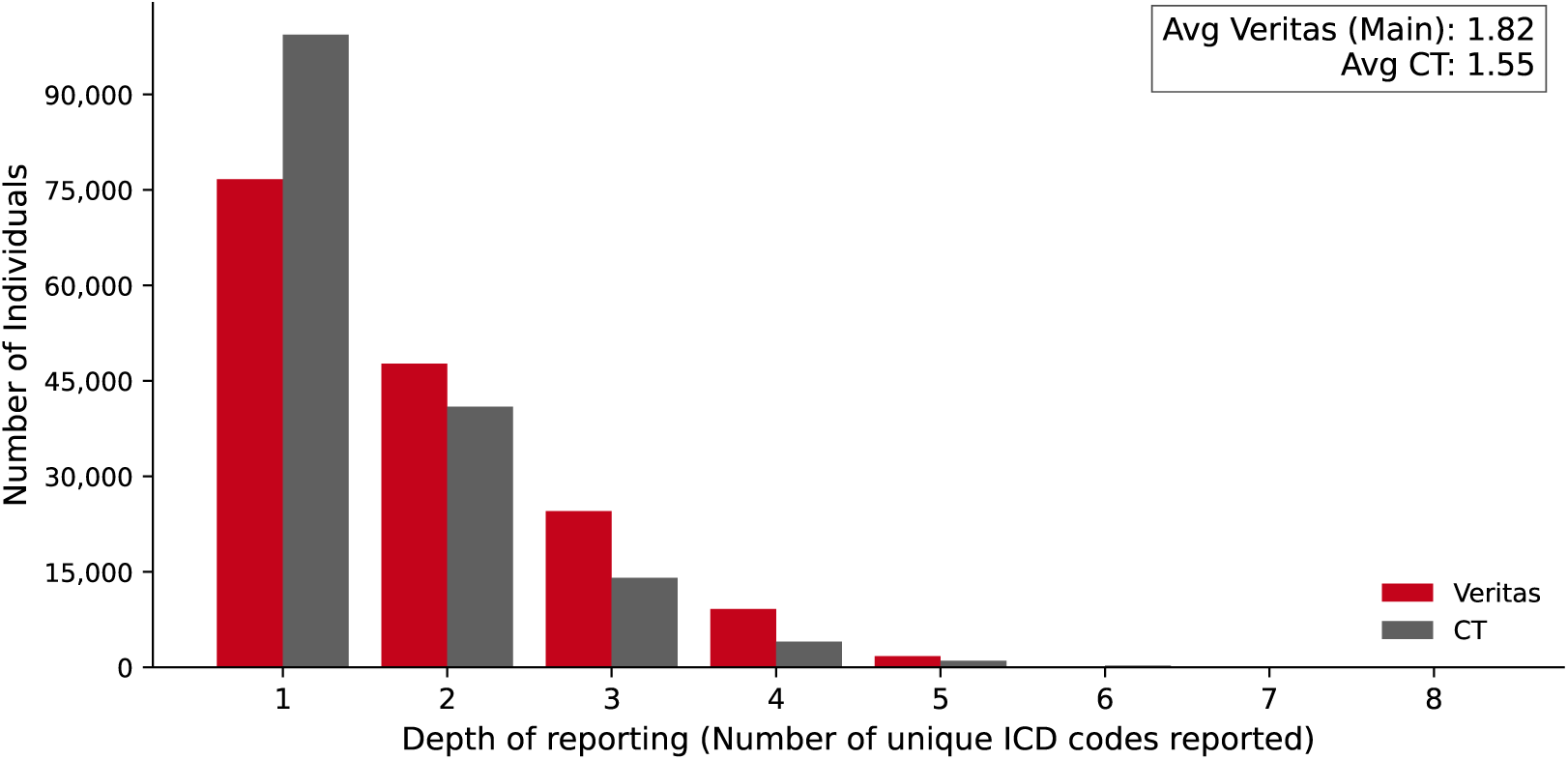
Depth of cause-of-death reporting within matched NCHS-113 groups. The distribution of distinct ICD-10-CM codes per individual is shown separately for Veritas and CT, restricted to individuals where both sources share at least one NCHS-113 group. Wilcoxon signed-rank test on non-zero differences: *n* = 57,810; proportion favouring Veritas = 77.7%; *p* ≈ 0.

#### 3.4.3 Specificity of Reporting

Among ICD-10-CM code pairs sharing the same three-character stem, Veritas more frequently assigned a more specific fourth-character code (i.e., a character other than ‘9’) than the CT death certificate. Results are presented in Figure 11.

**Figure 11.**
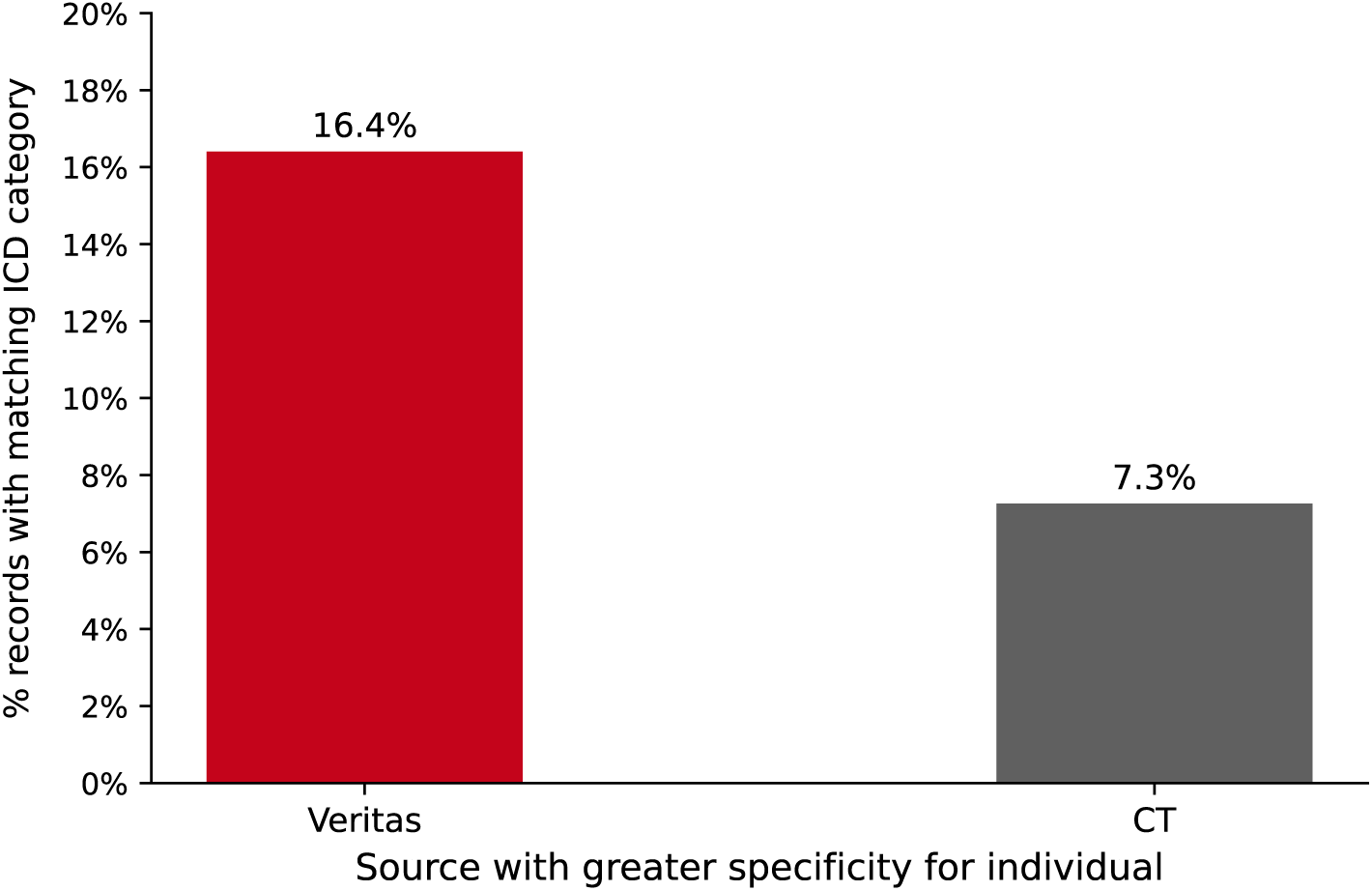
Specificity of ICD-10-CM code assignment among code pairs sharing the same three-character stem. Veritas more specific: Veritas fourth character ̸= ‘9’, CT fourth character = ‘9’. Equal: both sources in the same fourth-character specificity category. CT more specific: converse case.

## 4 Discussion

### 4.1 Metric Selection and Analytical Focus

The analytical framework centres on two complementary metrics: age-stratified chapter-level PPV and rank order stability (MARD). This pairing reflects the principal questions relevant to downstream deployment of the Veritas COD algorithm.

PPV is the most operationally relevant metric for applications in which Veritas output governs cohort membership or endpoint classification: pharmacovigilance signal detection, clinical trial outcome adjudication, and cause-specific mortality rate estimation.^23,24^ In these settings a false positive - assigning a chapter that is not present in the reference - introduces systematic inclusion error, and PPV directly quantifies that risk. Evaluation at the ICD-10-CM chapter level reflects the practical reality that most analytical applications aggregating across large populations are most interpretable at the chapter level: finer resolutions introduce inter-rater variability and coding specificity differences between sources that obscure the underlying diagnostic signal, while chapter-level agreement captures whether the algorithm and the death certificate are identifying the same fundamental disease domain.

MARD addresses the complementary question of rank fidelity. Applications that are sensitive to the primary versus contributing distinction - survival analysis endpoints, life insurance policy classification, actuarial models - require not only that Veritas detects the correct chapter but that it prioritises it appropriately. MARD quantifies rank instability for shared chapters, helping distinguish a rank misalignment between COD based on longitudinal claims and that listed on death certificates.

### 4.2 Age-Stratified Performance and Population Representativeness

The age-stratified PPV analysis reveals heterogeneity in algorithm performance across the mortality age spectrum that a population-aggregated metric would obscure. This heterogeneity has two principal sources.

The first is the epidemiological composition of mortality at different ages. The leading causes of death for individuals in the 25-44 age stratum - including substance use disorders, unintentional injury, and mental health conditions - involve clinical presentations whose evidence in medical claims is often indirect or episodic. By contrast, the leading causes in the 65+ stratum - cardiovascular disease, malignant neoplasms, chronic respiratory disease - generate sustained, high-acuity longitudinal claims signals that are more readily captured by the algorithm’s acuity-weighted scoring framework. In this space, we have demonstrated that longitudinal claims based COD assignment is a legitimate replacement for death certificate reporting.

The second source of age-dependent heterogeneity is insurance coverage density. Older individuals, particularly those enrolled in Medicare, carry longer and more complete longitudinal claims histories, providing the algorithm with a richer evidence base. This is the principal justification for the age ≥ 25 exclusion threshold applied uniformly to the cohort: the paucity of claims data for individuals under 25 would produce performance estimates that primarily reflect data availability rather than algorithm quality.

The marked right-skew of the study population toward older age groups was preserved throughout the analysis. Class reweighting or resampling to artificially balance age bands was not performed, as this would misrepresent the population to which the algorithm is applied in practice and distort macro-average PPV estimates by assigning equal influence to age bands that contribute very different numbers of deaths in the real-world mortality distribution. The power threshold functions as the appropriate mechanism for handling underpowered cells.

The overall validation conclusions are most robust for individuals aged 55 and above. This age range accounts for the majority of matched individuals and the majority of powered chapter-by-age-band cells. Results for the 25-34 and 35-44 bands should be treated as exploratory given the lower cell densities and wider confidence intervals in those strata.

### 4.3 Rank Concordance and Source Complementarity

The order analysis addresses whether, when Veritas and CT identify the same cause chapter, they assign it consistent rank positions. This question is distinct from detection concordance: it is possible for two sources to achieve high set overlap while exhibiting high MARD, indicating systematic disagreement about which cause is primary.

The interpretation of MARD should account for the well-documented unreliability of primary- cause designation on death certificates.^12,13^ Where both sources detect the same chapter but assign it different ranks, this may reflect a genuine disagreement about clinical priority, or it may reflect errors in the death certificate primary-cause selection. Per-chapter MARD values identify disease categories with the highest rank instability; for those chapters, supplementary investigation into the classification logic of both sources would be needed to attribute the instability to algorithm behaviour versus reference standard artefact.

A finding of high Jaccard with moderate MARD would suggest that the algorithm’s disease detection capability is strong and that ranking calibration is the primary area for development. A finding of low Jaccard with low MARD would suggest that the sources are drawing on systematically different clinical signals, and that breadth of detection is the primary gap. The mean rank comparison (Figure 7) provides a chapter-level view of directional rank bias, distinguishing chapters where Veritas systematically assigns higher or lower rank than the CT certificate.

### 4.4 Breadth, Depth, and the Structural Asymmetry of Longitudinal Claims Data

The breadth, depth, and specificity analyses are not concordance measures and should not be interpreted as evidence of algorithmic over-assignment. They characterise a structural asymmetry between the two source types: the Veritas algorithm draws on up to three years of longitudinal medical claims encompassing encounters across multiple clinical settings, provider types, and time windows, while the death certificate is completed at a single point in time, typically by a certifying clinician with limited access to the patient’s full clinical history. The observation that Veritas reports a broader, deeper, and more granularly specified set of causes is expected *a priori* and reflects the difference in observational scope rather than inflation of cause-of-death assignments.

The breadth finding (Veritas equal or greater in 76.8% of individuals) is consistent with the well-documented tendency for death certificates to under-report contributing causes.^23,24^ A single-event certificate captures the certifying physician’s assessment at the moment of completion; conditions that contributed to physiological decline over months or years-but were not acutely prominent at the time of death-are frequently omitted. Longitudinal claims records preserve evidence of these antecedent disease burdens, and the Veritas scoring framework is designed to weight both near-term acuity signals and longer-window recurrence patterns.

The depth finding (77.7% of non-zero differences favour Veritas; Wilcoxon *p* ≈ 0) indicates that within disease categories where both sources agree (matched NCHS-113 groups), Veritas identifies a finer-grained set of contributing ICD-10-CM diagnoses. Chronic disease processes recorded across many encounters naturally accumulate multiple diagnosis codes that a single death certificate entry for the same category would not capture.

The specificity finding indicates that Veritas more frequently assigns a specific, non-unspecified fourth-character ICD-10-CM code than the CT death certificate for codes sharing the same three-character stem. Specificity in ICD-10-CM coding depends on access to detailed clinical documentation: certifying physicians without access to the full medical record may default to unspecified subcategories, whereas algorithmic assignment can draw on procedure codes, modifiers, and encounter context that resolve specific subcategories.

Taken together, these analyses support the interpretation that longitudinal claims-based COD assignment provides a richer multi-dimensional characterisation of contributing disease processes than a point-in-time death certificate. This contextualises the PPV estimates in Sections 4.2 and 4.3: some apparent discordances between sources reflect the greater clinical depth of the Veritas output rather than algorithmic error.

### 4.5 Reference Standard Limitations and Performance Lower Bounds

A fundamental consideration in any death certificate validation study is that the reference standard itself is imperfect.^10,11^ Empirical studies consistently report a meaningful proportion of death certificates with incorrect primary cause attribution, with estimates typically ranging from 20% to 40% depending on the comparison method and clinical domain.^15^ An important consequence is that the PPV and other concordance metrics reported here are *conservative lower bounds* on true algorithm accuracy, not direct estimates of it.

This lower-bound property arises from the confusion matrix construction. For a given chapter *d*, a false positive is defined as a Veritas assignment that does not appear in the CT record. However, some fraction of these apparent false positives represent cases where Veritas has correctly identified a contributing cause that the certifying physician failed to record. Similarly, some apparent false negatives on the Veritas side reflect correct exclusions of causes erroneously included on the certificate. Since the true COD is unknown, these errors in the reference standard cannot be corrected analytically and are absorbed into the concordance statistics as algorithm errors.

The exclusion of R00-R99 chapter entries from the analysis reflects this lower-bound framing: symptom and sign codes are widely regarded as “ill-defined” in the cause-of-death literature,^9^ representing diagnostic imprecision in the reference standard rather than genuine underlying COD. Including them as valid reference entries would artificially inflate false-negative rates on the algorithm for clinically meaningful cause categories. Their exclusion therefore reduces one known source of reference standard noise from the concordance estimates, resulting in more interpretable PPV values.

### 4.6 Generalisability and Study Scope

The analytic cohort is restricted to individuals with a Connecticut death registration linked to Veritas RWD, processed to remove ICD chapters which will bias results. Generalisation of performance estimates to the full US mortality population across the more common causes of death should hold; though this is not explicitly tested due to absence of data. Note that RWD coverage rates, claims completeness, and the distribution of causes of death vary by state, insurer mix, and demographic composition. Future validation work extending to additional states or nationally representative samples would strengthen the generalisability of these findings.

## 5 Conclusions

This study presents a systematic external validation of the Veritas COD algorithm against an independent, state-level death certificate file, using a matched analytic cohort of *N* = 216,382 individuals (death years 2017-2025, age ≥ 25, V/Y chapter individuals excluded, R00-R99 entries removed from comparison), evaluated through two concordance metrics - age-stratified chapter-level PPV under the full-set concordance scenario, and rank order stability (MARD) for shared disease chapters - and three supplementary analyses characterising the completeness and granularity of COD reporting across sources.

The age-stratified PPV analysis reveals systematic heterogeneity in algorithm performance across the mortality age spectrum. Performance is most robust for individuals aged 55 and above, where both RWD claims density and chapter-by-age-band cell sizes are greatest. All reported PPV values represent conservative lower bounds on true accuracy, given the 20-40% error rate of the death certificate reference standard.^10,11^ The chapter level was selected as the appropriate taxonomic resolution for this analysis: it provides clinically meaningful disease domain groupings relevant to health analytics applications while permitting reliable cell-level PPV estimation after power-threshold filtering.

The order analysis (MARD) characterises the degree to which, for chapters identified by both sources, the two sources assign consistent rank positions. Chapters with elevated MARD represent cases where detection agreement exceeds ranking agreement; interpreting these values requires acknowledging that the death certificate primary-cause designation itself is unreliable in a non-trivial fraction of cases.

The completeness analyses demonstrate that Veritas reports equal or greater breadth of cause-of-death categories than the CT death certificate in 76.8% of individuals, with 77.7% of non-zero depth differences within matched NCHS-113 groups favouring Veritas (Wilcoxon *p* ≈ 0), and a greater frequency of specific fourth-character ICD-10-CM code assignments. These findings are structurally expected given the difference in observational scope between a three-year longitudinal claims record and a single point-in-time death certificate, and are consistent with the documented tendency for death certificates to under-report contributing causes.^23,24^

This study demonstrates that the Veritas COD algorithm produces a cause-of-death dataset with strong macro-level alignment with independent death certificate records, as evidenced by the ICD-10-CM chapter-level PPV analysis and the Jaccard set-overlap findings. Where Veritas and death certificate records diverge - in the cause categories identified, their rank priority, or their level of specificity - this divergence cannot conclusively be interpreted as evidence of algorithmic error. Given the well-characterised 20-40% primary-cause error rate of the death certificate reference standard,^10,11^ discordances at finer taxonomic levels are as likely to reflect genuine additional clinical signal captured from longitudinal claims data - or to identify cause-of-death attributions that are incorrect on the death certificate - as they are to represent algorithm deficiencies. The breadth, depth, and specificity findings reinforce this interpretation: three years of longitudinal claims data provide a richer characterisation of contributing disease processes than a point-in-time certificate can capture, and Veritas outputs at the ICD-10-CM code level represent a distinct and potentially more complete clinical record rather than a degraded approximation of the death certificate.

Applications in health economics and outcomes research (HEOR), retrospective cohort construction for real-world evidence, population-level mortality surveillance, and life sciences endpoint enrichment are well served by the macro-level alignment demonstrated here. More broadly, this study supports the conclusion that algorithmic cause-of-death ascertainment from longitudinal claims data constitutes a feasible and scalable alternative to death certificate-based attribution. It is not merely a proxy for the certificate: the structural depth of a multi-year longitudinal record captures disease burden that a point-in-time certificate cannot, and where the two sources diverge, the algorithm’s output represents a substantively richer characterisation rather than a discrepant one. Critically, large-scale comparison of algorithmically derived claims-based COD against death certificate records provides a principled and reproducible methodology for quantifying and characterising death certificate error rates at population scale - an objective that the existing literature has addressed only through small-sample clinical chart review studies with limited generalisability. The one domain where Veritas outputs require supplementary data is mechanism-of-injury attribution (ICD-10-CM chapters V00-Y89), where the structural absence of external-cause codes from standard claims feeds creates a coverage gap that longitudinal diagnostic data cannot bridge.

## Data Availability

All aggregated data produced in the present study are available upon reasonable request to the authors.

